# Brain magnetic resonance imaging software to support dementia diagnosis in routine clinical practice: a barrier to adoption study in the National Health Service (NHS) England

**DOI:** 10.1101/2024.08.02.24311223

**Authors:** Ludovica Griffanti, Florence Serres, Laura Cini, Jessica Walsh, Taylor Hanayik, Usama Pervaiz, Stephen Smith, Heidi Johansen-Berg, James Rose, Mamta Bajre

## Abstract

With the rise in numbers of people living with dementia and new disease modifying therapies entering the market, there is increasing need for brain magnetic resonance imaging (MRI) for diagnosis and safety monitoring. The number of scans that need reporting is expected to rapidly grow. Clinical radiology reports are currently largely qualitative and variable in structure and content. By contrast, research software typically uses automated methods to extract quantitative metrics from brain scans.

To better understand the unmet clinical need for brain reporting software for dementia we conducted a barrier to adoption study using the Lean Assessment Process (LAP)methodology. We first assessed the role of brain imaging in the diagnostic pathway for people with suspected dementia in the NHS in England. We then explored the views of (neuro)radiologists, neurologists and psychiatrists on the potential benefits and level of acceptance of software to support brain MRI analysis, using the FMRIB software library (FSL) as a technology exemplar.

The main perceived utilities of the proposed software were: increased diagnostic confidence; support for delivery of disease modifying therapies; and the possibility to compare individual results with population norms. In addition to assessment of global atrophy, hippocampal atrophy and white matter hyperintensities, additional user requirements included assessment of microbleeds, segmentation of multiple brain structures, clear information about the control population used for reference, and possibility to compare multiple scans. The main barriers to adoption related to the limited availability of 3T MRI scanners in the UK, integration into the clinical workflow, and the need to demonstrate cost-effectiveness. These findings will guide future technical development, clinical validation, and health economic evaluation.

## Introduction

In the UK, it is estimated that more than 944,000 people are currently living with dementia, with an associated economic cost forecast at £42 billion to the UK economy in 2024 (1) and predicted to increase to £94 billion by 2040 as the population ages (2)

NICE recommends the use of structural imaging, either Computerised Tomography (CT) or Magnetic Resonance Imaging (MRI) in the diagnosis of people with suspected dementia (3). Although CT scanning is widely available, cheaper, and as reliable as MRI in excluding potentially treatable conditions, MRI is more sensitive to detecting subtle changes, to assess vascular pathology, and to aid in differential diagnosis of dementia (4). Subtle brain changes can however be challenging to identify and quantify, as MRI brain scans are currently reviewed visually by a neuroradiologist. Various visual rating scales are available to enable a semi-quantitative assessment, but their impact and inter/intra-rater reliability is variable (5), and they require time and expertise (6). As a result, they have not been widely adopted into routine clinical practice, where the information included in imaging reports remains largely qualitative and of variable quality.

The 2021 National Audit for Dementia from the Royal College of Psychiatrists reported an increase in MRI scans from 26% of total neuroimaging scans performed in 2019 to 31.8% in 2021 (7), a figure set to increase dramatically in coming years as the population ages and new disease modifying therapies (DMTs) enter the market. Monoclonal antibodies are emerging DMTs for Alzheimer’s disease that require brain MR imaging for eligibility assessment and monitoring for amyloid-related imaging abnormalities (8). Extracting accurate quantitative measurements from MRI scans could aid in differential diagnosis for eligibility screening and longitudinal tracking of disease progression, treatment response and safety from side effects.

In contrast to the clinic, measures automatically extracted from brain scans to objectively assess atrophy and vascular damage are widely used in dementia *research*, with a variety of research software tools available (e.g. FSL, FreeSurfer, SPM, AFNI). Additionally, there are multiple commercial products emerging that are regulatory approved for clinical use (9). Despite this, there is limited adoption of dementia neuroimaging tools in clinical settings in UK clinics. Previously reported reasons for (neuro)radiologists not to perform quantitative evaluation in dementia imaging include lack of access to software algorithms and expertise, lack of picture archiving and communication system (PACS) integration, cost, users not comfortable with interpretation and the belief that it would be too time-consuming or user intensive (6). However, studies suggest that providing a quantitative analysis to clinicians improves interrater variability whilst increasing diagnostic confidence, sensitivity, and accuracy for diagnostic interpretation in dementia (10–12).

This barrier to adoption study aimed to assess the current neuroimaging approach used for diagnosing people with suspected dementia in the NHS England. We then explored the views of (neuro)radiologists, neurologists and psychiatrists on the potential benefits and level of acceptance of imaging analysis software in supporting MRI scan analysis. As a technology exemplar for the analysis software, we used the FMRIB software library (FSL) (13). FSL is a software package that includes over 300 tools capable of extracting various metrics from MRI scans, making it suitable for addressing this clinical need. The software is available for free for academic and personal use, and is licensed for commercial use. We asked the stakeholders perception of an FSL-based software with the following characteristics: 1) compatible with MRI scans and integrated in picture archiving and communication system (PACS); 2) automatically provides quantitative measurements of the volume of cerebral structures currently being measured as part of the dementia assessment (mainly global atrophy, hippocampal atrophy, and white matter hyperintensities); 3) displays data against healthy controls matched for age and sex and expressed as percentile of the normative reference population; 4) generates a quantitative report, prepopulated with hospital number sets, age, date of birth, date of scan and measurements for cerebral structures measured in clinical diagnosis and assessment of dementia.

## Material and methods

The Health Innovations Oxford and Thames Valley and Wellcome Centre for Integrative Neuroimaging (the developers of FSL) sought to explore how an imaging analysis software tool could support dementia diagnosis in routine NHS clinical practice.

The Lean Assessment Process methodology (14) is used to align evidence generation with available resources at an early stage to explore the unmet clinical needs and the potential barriers to adoption of a new product. The methodology assesses the feasibility of implementing a technology in a care pathway using a preliminary assessment of design, value, and evidence reliability (15). LAP was used to evaluate how the FSL-based software capability and design aligned with the unmet needs in the current dementia diagnosis pathway as well as to identify any potential barrier to its adoption in a clinical setting by engaging with key stakeholders early on in the product development process. Briefly, the dementia diagnosis care pathway was mapped using published guidelines. Ten key stakeholders to interview were identified through both literature review and recommendations. All interviewees were healthcare professionals working in relevant roles within the dementia care pathway across 8 different NHS Trusts in England and included neuroradiologists, general radiologists, FGD-PET radiologists, neurologists and Old Age psychiatrists working in memory clinics. Documents describing the technology were circulated to stakeholders prior to engaging with them. Questionnaires used during the interview contained both qualitative questions on the current landscape and current clinical practices, quantitative product perception questions using a 7-point scale as well as human factor tools adapted from the perceived usefulness questionnaire (16), the stakeholders questionnaire (17) and the net promoter score ((18), which predicts market potential. Qualitative and quantitative data were thematically analysed to build an understanding of the technology acceptability, perceived usefulness and likelihood of its adoption by stakeholders. Moreover, user preferences, stakeholder acceptance, product design and potential barriers of its adoption in the NHS in England were evaluated to further refine the value proposition.

## Results

### Views on the current landscape

NICE guidelines were largely followed by stakeholders. The current neuroimaging referral pathway is summarised in Fig 1.

**Fig 1.**
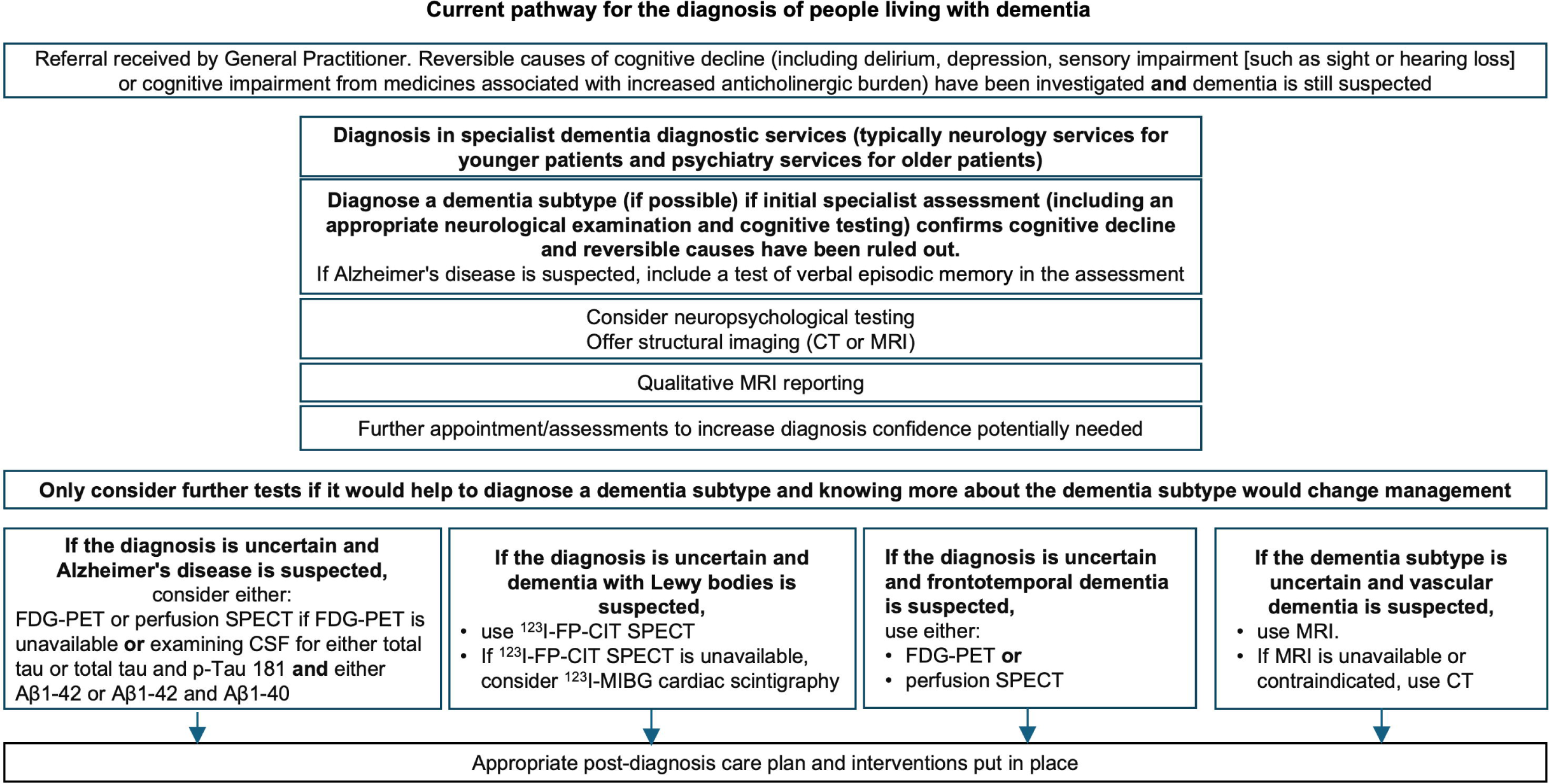
Current pathway for the diagnosis of people with suspected dementia in the NHS England (NICE guidelines [NG97] Dementia: assessment, management and support for people living with dementia and their carers). Legend: CT: computed tomography, MRI: Magnetic Resonance Imaging, FDG-PET: fluorodeoxyglucose-positron emission tomography-CT, SPECT : single-photon emission CT, CSF: Cerebrospinal fluid, P-tau: phosphorylated-tau, Aβ: β-amyloid, pTau: phosphorylated tau, ESNR: European Society of Neuroradiology.

It was recognised by stakeholders that the number of patients referred for neuroimaging is variable across Trusts and clinicians. Most Trusts have a cut-off age to determine whether a patient should be seen by the cognitive neurology team (younger patients) or by an Old Age psychiatrist (older patients). Patients referred to neuroimaging through neurology would all receive an MRI, as the clinicians will be looking for potentially subtle and harder to detect changes in the hippocampus. For psychiatry referrals, CT was predominantly used to rule out alternative causes of cognitive impairment, and is well tolerated in older adults. The level of agreement amongst stakeholders on whether MRI is better than CT in the dementia diagnosis pathway was 88.6%, with clinicians commenting that although the pattern of atrophy or the degree of small vessel disease can be seen on CT, it is “*not as good as what you can assess with the MRI”*. However, access to MR scanners was limited.

Establishing the diagnosis lies with the referring clinicians with the support of neuroimaging. In some cases, further imaging assessments are required (Fig 1). However, in practice, this concerns a very small number of patients, judiciously selected, as to not overwhelm the radiology services which was commented upon as being *“exceptionally limited and often available off-site in another larger regional hospital”*.

When discussing reporting time, radiologists mentioned that it would take them about 10-15 minutes to review a scan. Neurologists explained that they typically receive the report back from radiology within a few days, whereas for psychiatrists, turnaround times varied across Trusts, ranging from around 2 weeks, up to 6-12 weeks, with up to 3 months from the point of decision to refer to a scan to the person coming back to the clinic. The delay was not necessarily due to radiology, but also to the difficulty of finding an available scanner to perform imaging or a time slot at the memory clinic for reviewing the patient. In the psychiatrists’ opinion, the main adverse effect of the delay related to the fact that patients were anxious to receive a diagnosis but that the delay did not really affect patients’ management as current treatment options for dementia are limited. Stakeholders also mentioned that reporting was sometimes outsourced to external radiologists to decrease delays when the capacity of Trust’s radiology services was limited.

Clinicians would provide the radiology department with a short clinical history of the patient, performance on cognitive assessments and the suspected patient’s diagnosis on the referral letter. Radiologists commented that they find it particularly helpful when a good clinical history is provided. For example, the presence of “*word finding difficulties and aphasia*”, or of “*parkinsonian aspects associated with hallucinations potentially indicative of Lewy bodies dementia*” would guide them. Radiologists commented that conversely, referrals with requests such as “*please, is there hippocampal atrophy*” or “*memory loss*” without any details were found particularly unhelpful.

Most of the radiologists used their own reporting templates that they would manually populate with measurements and scores. Vascular burden and microbleeds were assessed before measuring the pattern of atrophy. The decision to assess specific areas would be guided by the referral letter. Some of the cerebral regions assessed by the radiologists interviewed included medial temporal areas, cingulate region, the dorsal regions, convexity of the palate, frontal lobes, cerebellum, mamillary bodies, corpus callosum and brain stem. (Neuro)radiologists would sometimes provide quantitative assessments using visual rating scales or a description of severity. Neuroradiologists mentioned reporting MTA scores, ERICA scores, KOEDAM scores, vascular burden as mild, moderate, severe and FAZEKAS scores.

Unlike the neurologists, none of the psychiatrists interviewed had access to the scan images, so they would rely purely on the radiologists’ reports. All stakeholders emphasised that neuroimaging was part of the diagnostic assessment but not a standalone diagnostic tool. Overall, the content and quality of reports that clinicians received from radiology were hugely variable.

Stakeholders agreed on the need for training and better-quality reporting of MRI scans in dementia diagnosis. The level of agreement on the need for software to support the analysis of volumetric MRI scan and provide quantitative data was 80.7%.

### Perception of the technology

Table 1 summarises the answers of the stakeholders to questions about their individual perspective (level of agreement) and the perceived usefulness of the technology. 83.3% of stakeholders stated they would promote the technology in future. When asked about the potential added value, impact on clinical practice and key functionalities of the technology the stakeholders discussed several aspects.

**Table 1.**
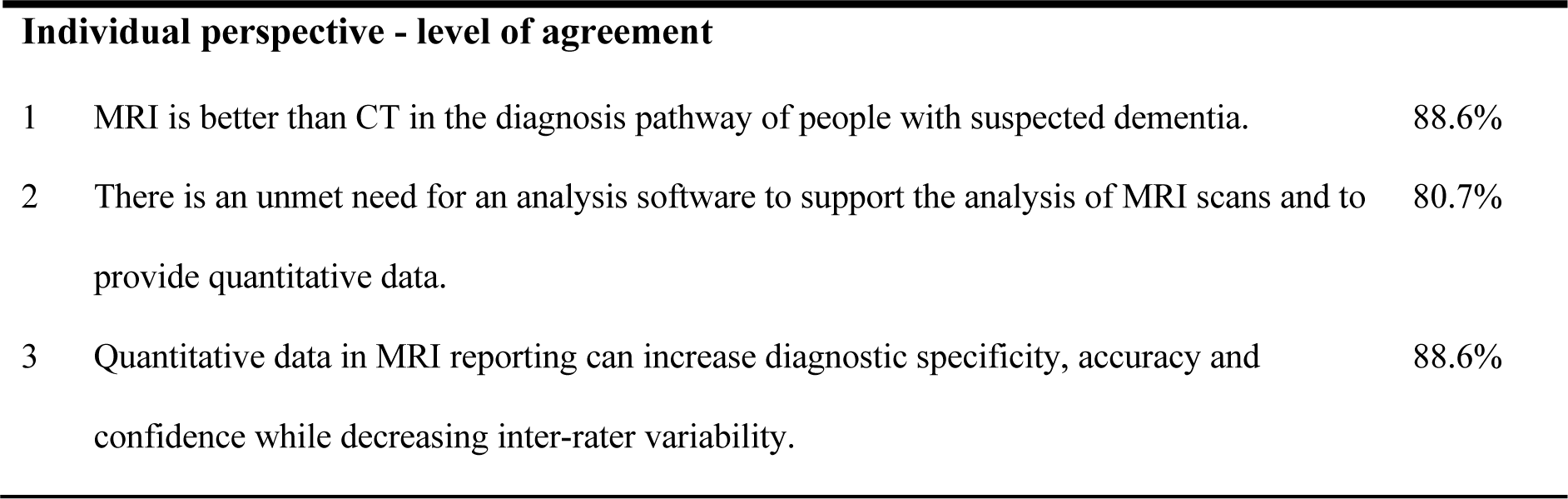
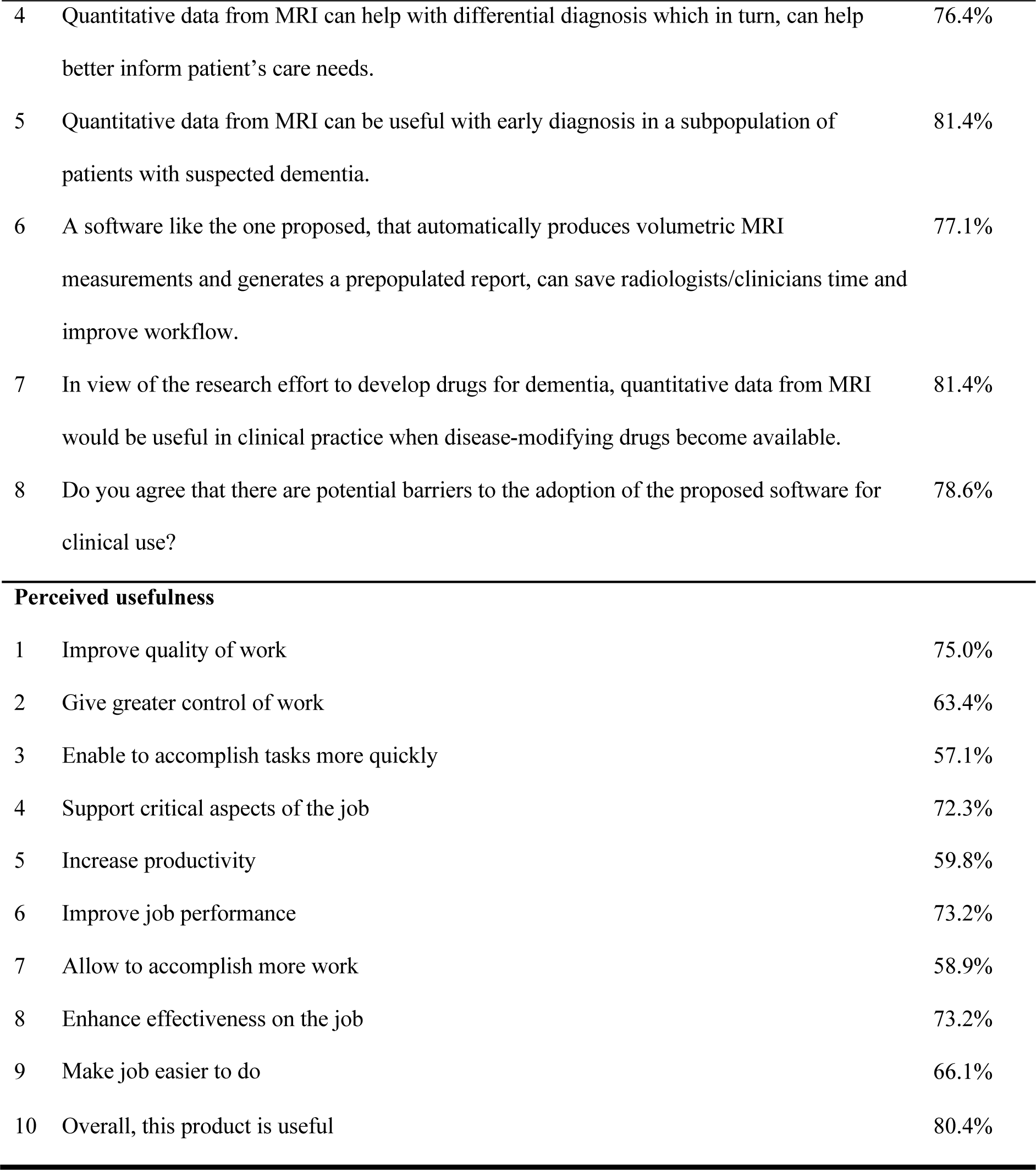
Stakeholder’s average level of agreement and perceived usefulness of the proposed analysis software based on FSL.

| Individual perspective - level of agreement |  |  |
| --- | --- | --- |
| 1 | MRI is better than CT in the diagnosis pathway of people with suspected dementia. | 88.6% |
| 2 | There is an unmet need for an analysis software to support the analysis of MRI scans and to provide quantitative data. | 80.7% |
| 3 | Quantitative data in MRI reporting can increase diagnostic specificity, accuracy and confidence while decreasing inter-rater variability. | 88.6% |
| 4 | Quantitative data from MRI can help with differential diagnosis which in turn, can help better inform patient's care needs. | 76.4% |
| 5 | Quantitative data from MRI can be useful with early diagnosis in a subpopulation of patients with suspected dementia. | 81.4% |
| 6 | A software like the one proposed, that automatically produces volumetric MRI measurements and generates a prepopulated report, can save radiologists/clinicians time and improve workflow. | 77.1% |
| 7 | In view of the research effort to develop drugs for dementia, quantitative data from MRI would be useful in clinical practice when disease-modifying drugs become available. | 81.4% |
| 8 | Do you agree that there are potential barriers to the adoption of the proposed software for clinical use? | 78.6% |
| Perceived usefulness |  |  |
| 1 | Improve quality of work | 75.0% |
| 2 | Give greater control of work | 63.4% |
| 3 | Enable to accomplish tasks more quickly | 57.1% |
| 4 | Support critical aspects of the job | 72.3% |
| 5 | Increase productivity | 59.8% |
| 6 | Improve job performance | 73.2% |
| 7 | Allow to accomplish more work | 58.9% |
| 8 | Enhance effectiveness on the job | 73.2% |
| 9 | Make job easier to do | 66.1% |
| 10 | Overall, this product is useful | 80.4% |

#### Increased confidence

Both radiologists and clinicians mentioned increased confidence in diagnosis as the main added value. The level of agreement amongst stakeholders on the capability of quantitative MRI reports to increase diagnostic specificity, accuracy and confidence while decreasing inter-rater variability was 88.6%. Having quantitative data would give them greater confidence in the MRI findings and potentially reduce the need for further FDG-PET scans, which are more expensive and with less capacity. However, some neuroradiologists raised concerns about people with little experience of dementia imaging using the sort of software that might give a sense of “false reassurance to a general radiologist”.

> *“Then you can very, very quickly say: okay the pattern in this convincingly matches the clinical input. Nothing else to be said. You move on. So it would be straight forward for the patient, straight forward, for the clinician, straight forward for radiologist. So you have less patients in the limbo they are until you reach a diagnosis, which sometimes is very time consuming.”-* Consultant neuroradiologist

> *“I think a neuroradiologist would know what is going on but if you don’t have a neuro background, and it is just purely imaging, that is a different conversation”.* - Consultant neuroradiologist

#### Support delivery of disease modifying therapies (DMTs)

The level of agreement on whether quantitative MRI would be useful to clinicians when DMTs become available was 81.4%.

Some stakeholders believed MRI in this pathway will be used to look for dementia drugs contraindications because of the risk of bleeds associated with their use. Others felt that MRI will be used to monitor the effects of these drugs on disease progression and in that context, quantitative MRI will provide valuable data. However, stakeholders also commented that the whole dementia diagnosis pathway is likely to change with the arrival of dementia-modifying drugs and that eligibility criteria for these expensive treatments will be based on fluid biomarkers such as serum amyloid levels or amyloid- PET rather than MRI. In some stakeholders’ views, the future of MRI in the dementia pathway will not be for diagnostic purposes as fluid biomarkers will likely have higher predictive value, lower cost and higher accessibility than MRI. Instead, they see MRI being used for safety monitoring and for determining which treatments patients should receive.

> “I would say vital. So when we get to dementia modifying drugs, there will be a pathway. I think the pathway will involve both neuroimaging, potentially blood biomarkers for negatives and then only in the people where you’ve really done everything to rule out other causes, do you do the lumbar puncture.” - Consultant psychiatrist

> *“…[diagnostic] criteria is going to be amyloid status rather than any other parameters, so it would be amyloid-PET or hopefully serum levels of amyloid and other relevant actors” -* Consultant psychiatrist

> *"…the future of MR imaging in dementia diagnosis is not going to be diagnostic. It’s going to be safety for treatment … The diagnosis will be done in a different way.” -* Consultant neurologist

#### Comparison with a control population

One of the main perceived benefits of such a software tool was that it would provide comparison of the patient’s measurement with those of a normative population matched for age and sex. All stakeholders commented that this information would be very valuable both in a younger population of patients where the volume of cerebral structures such as the parietal cortex is highly variable, as well as in the older population where the rate of atrophy varies significantly between individuals and does not necessarily correlate with cognitive impairments. However, stakeholders were keen to be provided with information on the composition of the control population used to display the results (and train the AI if applicable). Questions asked pertained to how many control scans were going to be used (to judge the variance of the results), where the control population came from (to make sure it is representative of the population where the tool is being deployed), and if an amyloid scan has been performed on these healthy individuals (to check whether they could be in a pre-dementia state). Uncertainty around the control population was also the main reason for loss of confidence in commercially available software that had been tried by some stakeholders.

#### Assessment of microbleeds and vascular pathology

Microbleeds were seen as extremely important to consider. Although their assessment is not part of the current dementia diagnosis, stakeholders felt that they will need to be measured more accurately by neuroradiologists when new dementia drugs are available. Quantification of microbleeds and vascular change would inform on what the bleeding risk would be for disease management and informing treatment.

> *“At the moment, we don’t use and I don’t need it, but I can see that a rapid automated assessment to determine the risk of bleeds and that would become of particular value when we come to the treatment for the amyloid which is not very far away.” –* Consultant neuroradiologist

Other added values of the technology for stakeholders were the possibility to detect subtle changes, save time saving, and differentiate age related changes from pathological changes. Other key functionalities mentioned to be added to the software were comparison of multiple scans, and segmentation of multiple brain structures to assess atrophy patterns and other abnormalities.

### Barriers to adoption

Stakeholders recognised the likelihood of barriers to the adoption of a software tool like the proposed one in the clinical setting, with 78.6% agreement to the question: "Do you agree that there are potential barriers to the adoption of the proposed software for clinical use?"

Key barriers to adoption of such a software tool were the limited access to MRI scanners, in particular 3 Tesla, in NHS services and therefore CT is widely relied on in the dementia care pathway. The psychiatrists interviewed estimated the ratio to be 2/3 CT and 1/3 MR in one trust and 95% of CT in the other. The demand for MRI scans for eligibility screening and safety monitoring in the dementia care pathway will increase in the coming years when DMTs come to market. However, in the UK there are currently 8.6 MRI scanners per million people compared to 38 and 35.3 in the US and Germany respectively (19).

> *“MRI access is an issue. We are not really encouraged to ask for MRI scans. So if I asked for CT scan, they’re pretty much will always say yes; If I ask for an MRI scan, they’ll triage it much more carefully.”-*

#### Consultant Psychiatrist

> *“Because of various things, as we’ve discussed, about capacity, availability, some patients can’t have an MRI. I think in an ideal world, you would want everyone to have an MRI, but we don’t live in an ideal world.”-* Head and Neck Consultant Radiologist

Additional barriers identified by stakeholders were related to integration in the clinical workflow (IT and time requirements); structural issues within the NHS making it hard for innovations to be adopted; potential issues with information governance; managers’ attitudes towards innovation; and the cost of the technology. Stakeholders explained that to introduce an innovation into the department, one needs to write a business case to demonstrate that the innovation will be cost-effective. For example, *“to demonstrate that saving time on reporting scan would result in xx extra scan reported, equating to ‘x’ amount of savings”*. This business case would therefore be highly dependent on the number of dementia scans processed in the radiology department. If dementia scans are not the bulk of their activity, radiologists will have to demonstrate that the software could be used for imaging of other pathologies, such as multiple sclerosis. However, stakeholders also thought that although cost would be an issue, they were confident that there were ways around it, for example using champions to promote the analysis software.

## Discussion

The current study aimed at exploring the perceived usefulness and level of acceptance of a brain imaging software tool, using FSL as a technology exemplar for the analysis software, in providing quantitative reporting of MRI scans in the dementia diagnosis pathway. To gain a broad view of potential barriers to adoption in clinical practice in NHS England, we interviewed stakeholders who worked in 8 different Trusts across England in relevant roles in the dementia pathway.

Although imaging is an integral part of the dementia pathway in most cases, the choice of CT vs MRI is made mainly based on the referring service (CT for psychiatry, MRI for neurology) and access to MRI scanners. Reporting time is heterogeneous for psychiatry and neurology, and the content of reports is variable. Reports usually contain common elements, but the level of detail provided, as well as the use of visual rating scales is not consistent. Despite clinicians and radiologists agreeing on the usefulness of structured reports, there was no consensus on a common template. This is in line with what has been recently reported by the European Society of Radiology: while in many countries national radiological societies have launched initiatives to further promote structured reporting, cross-institutional applications of report templates (e.g., for registries or research) and incentives for usage of structured reporting (monetary or structural) are lacking (20). In addition, the specificity in the referral question, the specialty of the radiologist/neuroradiologist, and specialty of the referring clinician seem to contribute to the heterogeneity in content as well as to the level of trust in the report. Importantly, while neurologists have access to the images and often personally review them alongside the radiology report, psychiatrists typically only have access to the report and not the images.

In this context, stakeholders agreed that there was a need for software to provide volumetric quantification of brain MRI and for better reporting in the dementia diagnosis pathway. With the shortage of radiology workforce (29% in 2022 (21)) and the predicted increased number of referrals for dementia diagnosis scans, the proposed software was seen as a useful radiology support tool for analysing brain scans. For radiologists, although the software was perceived as not saving time per se, they would expect it to increase their diagnostic confidence by allowing patients to be compared to normative data. For clinicians, the creation of a structured report collating all the information needed to make a diagnosis was also seen as helpful. More importantly, it could reduce the variability between reporters and decrease the subjectivity of the report, a point seen as important by clinicians in the context of shortage of neuroradiologists. If the proposed FSL-based software was coupled with additional features such as measuring other cerebral structures, comparing scans and microbleeds measurement, this software could serve several purposes within the dementia pathway: as a diagnostic tool to better identify the subtype of dementia and reduce the need for further investigation; as a monitoring tool to measure subtle changes and evaluate disease progression or the effect of disease-modifying treatments; and/or as a tool to identify risk of bleeding, particularly relevant in view of the safety monitoring required for dementia drugs soon to be available.

When discussing their confidence in the outputs of the software, stakeholders explained that they would have to be provided with information on the software’s sensitivity and specificity. Information on the control population used would be useful to reassure users that it is representative of the local population both in its diversity and across all ages. Stakeholders also mentioned the important balance between the software increasing confidence and the risk of creating a “false sense of reassurance” that the measurements are adequate if they lack the knowledge to quality-control the findings of the software.

One of the main barriers to adoption of such software was the limited access to MRI scanners, and therefore the fact that CT is widely relied on in the dementia care pathway. Other barriers mentioned by all stakeholders were the cost and the need to demonstrate that the software would be a cost-effective tool for brain reporting. Stakeholders were unsure whether using quantitative MRI could save them time or reduce the need for further investigations. Some stakeholders were of the view that the role of quantitative MRI in the future dementia diagnosis pathway would only be to assess the risk of bleeds, while biofluid measurements will probably become the mainstream diagnostic biomarkers for early dementia. Showing that the software could be useful in other pathways, and finding champions to promote it, were proposed by stakeholders as potential solutions to convince NHS managers to acquire it.

While covering a broad complement of stakeholders in relevant roles in the dementia diagnosis pathway, across 8 different NHS trusts, the stakeholder sample size was small (n=10) which is a limitation of this study.

Altogether, this barrier to adoption study using LAP methodology reported that stakeholders were very positive about the potential usefulness of brain analysis software tool in the dementia diagnosis pathway, as reflected by the overall usefulness score of 80.4%, a score even greater for radiologists as primary users of the software.

This study highlighted several future directions towards the adoption of brain imaging software to support dementia diagnosis in the NHS England. Regarding the software technical requirements, specific measures (microbleeds, segmentation of multiple structures) and functionalities (representative control population, comparison of scans over time) were mentioned as important to include to meet clinical needs. These improvements should be supported with technical validation studies to improve confidence in the software. A clinical evaluation study involving clinical end-users would test the value of the software compared to a standard reading. This could include measuring the reporting time and accuracy, assessing confidence in diagnosis, as well as gathering feedback around the usability and acceptability of the software. An integration test in a clinical setting would enable assessing the impact of the software in the clinical workflow. Joint clinical-research facilities like the Oxford Brain Health Clinic (22) provide an ideal translational interface to test the software on data from real-world clinical populations in a research setting, while building evidence for further adoption. Finally, a real-world health economic evaluation will be required to determine the cost-effectiveness and to capture the clinical effectiveness as well as gathering further clinical evidence.

## Data Availability

Anonymised data produced in the present study are available upon reasonable request to the authors for non-commercial purposes.

## Notes

### Competing Interest Statement

SS and LG receive royalties from licensing of FSL to non-academic, commercial parties. SS is part-owner of a neuroimaging analysis company SBGneuro. The remaining authors report no potential conflicts of interest.

### Funding Statement

This project was supported by the Wellcome Centre for Integrative Neuroimaging (203139/Z/16/Z, 203139/A/16/Z and 215573/Z/19/Z) and by the NIHR Oxford Health Biomedical Research Centre (NIHR203316). LG was also supported by an Alzheimer's Association Grant (AARF-21-846366). The views expressed are those of the author(s) and not necessarily those of the NIHR or the Department of Health and Social Care. For the purpose of open access, the author has applied a CC BY public copyright licence to any Author Accepted Manuscript version arising from this submission.

### Author Declarations

The project was conducted by Health Innovation Oxford and Thames Valley, hosted by Oxford University Hospitals NHS Foundation Trust. This work does not require approval from an NHS Research Ethics Committee, according to the following criteria: participants in the study were not randomised to different groups, the study does not demand changing treatment/patient care from accepted standards, and the findings are not going to be generalisable.

